# Molecular Imaging of Systemic Inflammatory Response in Lymphatic Organs Provides Prognostic Value after Re-Perfused Acute Myocardial Infarction

**DOI:** 10.1101/2025.09.04.25334995

**Authors:** Theresa Reiter, Anna-Lena Dörrler, Natalie Hasenauer, Sebastian E. Serfling, Takahiro Higuchi, Nils Kraus, Wolfgang R. Bauer, Willibald Hochholzer, Gustavo Ramos, Samuel Samnick, Ulrich Hofmann, Stefan Frantz, Andreas K. Buck, Aleksander Kosmala, Rudolf A. Werner

## Abstract

**Background:** Acute myocardial infarction (AMI) triggers local inflammation in the injured myocardium, followed by a systemic inflammatory response of lymphatic organs. This prospective trial investigated whether molecular imaging of lymphatic organs (spleen, bone marrow, and heart-draining lymph nodes (LN)) identifies individuals predisposed to functional recovery during follow-up after AMI.

**Methods:** 41 timely re-perfused ST-elevation AMI patients received positron emission tomography (PET) using ^68^Ga-PentixaFor, a radiotracer targeting C-X-C motif chemokine receptor 4 (CXCR4) post-AMI. Patients were allocated to an early (2-4 days) or late PET imaging group (5-8 d) based on tracer availability. To determine left ventricular ejection fraction (LVEF) and infarct size, cardiac magnetic resonance imaging was conducted at baseline and repeated after six (follow-up (FU) 1, available in 38/41) and twelve months (FU 2, 36/41). As endpoint, an LVEF increase of ≥5% relative to baseline was then defined as short- (at FU 1) and long-term functional (at FU 2) recovery. We also determined association of ^68^Ga-PentixaFor uptake in the infarct territory and lymphatic organs with outcome relative to established clinical and imaging biomarkers.

**Results:** At FU 1, functional recovery was recorded in 21/38 patients. Univariate analysis identified baseline LVEF (Odds Ratio (OR), 0.80, P=0.002) and uptake derived from heart-draining LN (OR, 0.21, P=0.03) as predictor of functional recovery, while only LVEF reached significance at multivariate analysis (OR, 0.72, P=0.007). At FU 2, functional recovery was observed in 21/36 patients and LVEF (OR, 0.84, P=0.005) and splenic PET signal (OR, 2.46, P=0.04) provided prognostic value at univariate analysis. Both parameters remained significant at multivariate outcome analysis (LVEF: OR, 0.73, P=0.01; spleen: OR, 4.17, P=0.04), indicating that low baseline LVEF and high splenic tracer uptake are prognostic for improved long-term functional outcome. Established risk factors of cardiac damage (infarct size) or inflammation (C-reactive protein, white blood cell counts) failed to reach significance for functional recovery at FU 1 and FU 2.

**Conclusions:** Along with baseline EF, imaging of chemokine receptor expression in the spleen provides a complementary in-vivo biomarker for long-term functional recovery. Whole-body PET-based assessment of systemic inflammatory response in lymphatic organs may therefore open avenues for immune modulation strategies in patients after AMI.

## INTRODUCTION

Recent years have witnessed an increasing amount of evidence for immune modulating therapeutic interventions after myocardial infarction (MI) ^1-3^. However, there is an unmet need in biomarker or imaging-guided approaches for clinical decision-making on immune-modulatory therapy. Laboratory biomarkers of inflammation including white blood cell count (WBC) or C-reactive protein (CRP) can identify high-risk individuals prone to cardiovascular events ^4^ or to guide targeted therapeutic interventions ^3^, but are not suited to identify inflammatory changes on a cellular level in certain organs of interest ^5^. Cardiac magnetic resonance imaging (CMR) is the method of choice to determine the current cardiac functional status and multiple trials have reported on respective associations between CMR-derived parameters and outcome ^6,7^. Despite those clinical benefits, CMR has limited specificity for detecting inflammation in the heart. Moreover, MR-imaging of immune-activity in secondary lymphatic organs has not been established. The tracer principle, however, goes beyond morphological assessment of function and allows for evaluating the underlying pathophysiology at the target site (infarcted myocardium) and remote organs, which are involved in the systemic immune response ^5^.

Among others, targeting C-X-C motif chemokine receptor 4 (CXCR4) is crucially involved in the tight orchestration of recruiting and homing of stem cells to the under-perfused cardiomyocytes after acute MI (AMI) ^8^. In the injured heart, CXCR4 is not confined to one cell type, but lymphocytes account for the vast majority of CXCR4-positive cells in lymph nodes (LN, >90%) ^9^. The broad expression of CXCR4 in bone marrow-associated progenitor cells or natural killer cells also provides a rationale for monitoring chemokine receptor expression in the spleen and bone marrow after an acute event ^10,11^. Due to its considerable low cardiac background activity, ^68^Ga-PentixaFor, a radiotracer targeting CXCR4 on a broad spectrum of leukocytes, has been investigated in recent years in varying pre- and clinical scenarios post-MI ^12^. Of note, as radiotracers are applied systemically, chemokine receptor expression of organs of interest including heart-draining LN, spleen and bone marrow can be determined through a whole-body positron emission tomography / computed tomography (PET/CT) approach. In this regard, uptake in lymphatic organs including splenic in-vivo CXCR4 upregulation has already provided prognostic value in other clinical scenarios ^13^.

As such, we hypothesized that imaging of CXCR4 expression in organs involved in the systemic immune response can provide incremental prognostic performance relative to other established clinical parameters for functional recovery in first AMI patients. This may allow to determine high-risk individuals prone to cardiac functional changes during short- and long-term follow-up (FU).

## MATERIAL and METHODS

### Study Design and Participants

This prospective trial enrolled 41 patients (12 females, 59±9 years) which received CXCR4-directed ^68^Ga-PentixaFor within 8 days after the acute ST-elevation MI, allocated to an early (2-4 days) or late imaging group (5-8 d) based on tracer availability. Further inclusion criteria included minimum age of 18 years and immediate catheterization, along with stable clinical course. Exclusion criteria included hemodynamic instability > 48 h after immediate catheterization, known coronary or structural heart disease, multivessel disease, non-ST-elevation MI, active cardiac or ferromagnetic implants, sarcoidosis, acute inflammatory disease, contraindications for CMR imaging, impaired kidney function, immunosuppression, pregnancy or breast feeding. Patients also received CMR at baseline after AMI, followed by repeated CMR at six (FU 1) and twelve months (FU 2) to determine early and long-term functional recovery. We recorded standard laboratory values of cardiac damage, including peak Troponin T (in ng/ml), peak creatine kinase (CK, in U/l), peak lactate dehydrogenase (LDH, in U/l), and N-terminal prohormone of brain natriuretic peptide (NT-proBNP, in pg/ml). At day of PET imaging, creatinine (in mg/dl) and systemic inflammatory markers, including WBC (in x1000/µl) and CRP (in mg/dl), were also collected from the blood. Patients gave written informed consent prior to the study and the local ethical committee approved this trial (#192/21). Details on patient characteristics can be found in **Supplementary Tables 1 and 2**. ^68^

### Imaging and Outcome

Ga-PentixaFor was produced in-house ^14^, following good manufacturing practice as described in by using a Scintomics synthesis module (Gräfelfing, Germany) and single-use cassette kits provided by ABX (Radeberg, Germany). Image acquisition took place 45 minutes after injection of 145±13 MBq ^68^Ga-PentixaFor, with a field-of-view ranging from the cervical region to the spleen. In 3D mode, measurements were taken for 2 minutes per bed position, and image reconstruction was performed using an ordered subset expectation maximization algorithm (attenuation-weighted ordered subsets expectation maximization algorithm; 4 iterations, 8 subsets). Static PET images over 20 min were conducted on either a Biograph mCT 64 or 128 PET/CT. Guided by PET/CT, we set volumes of interest on the PET portion in the infarcted and remote myocardium, spleen, bone marrow of the vertebral bodies and heart-draining and remote LN in the axillary region (with no discernible tracer uptake) to determine peak standardized uptake values (SUV_peak_) ^12^. As described previously ^15^, we then calculated the uptake ratio of infarcted/remote myocardium (referred to as infarct) and heart-draining/remote LN. For the latter, we then determined the median ratios of all investigated LNs (referred to as LN). Low-dose CT was also conducted for attenuation correction.

All CMR scans were performed on a 3.0T clinical MRI scanner (Achieva DS, Philips Healthcare, Best, The Netherlands) and performed following current recommendations ^16^. In short, the protocol included cine imaging in standard short and long axes cine imaging, T2 weighted edema imaging, quantitative T1 and T2 mapping as well as late gadolinium enhancement (LGE) imaging. The latter was performed 10-15 min after application of Gadobutrol 0.01 mmol/kg (Bayer Vital GmbH, Germany). For image analysis, the endomyocardial border in both diastole and systole were traced manually in order to calculate left ventricular ejection fraction (LVEF). The LGE short axis stack was also segmented manually. Scar tissue was identified as a deviation in signal intensity by 5 standard deviations or more from the signal intensity of remote regional tissue, and LV mass, LGE mass and LGE/mass (in %, reflecting infarct size) were calculated. Patients received CMR at baseline after AMI, followed by repeated CMR at six (FU 1) and twelve months (FU 2) to determine an LVEF increase of ≥5% as the endpoint of the study (defined as long- and short-term functional recovery) ^17^.

### Statistics

We used R (version 4.3.2, R Core Team, 2023, Vienna, Austria) with package rms (6.7.0) and Prism (version 10.4.2, GraphPad, San Diego, California). Values are presented as mean ± standard deviation or range in parentheses. Student’s *t* test was used to test for differences between LVEF at baseline relative to follow-up. Uni- and multivariate Cox regression analyses were applied to determine baseline parameters with prognostic values (15), with time-point of PET imaging serving as moderating variable. Using QQ plots, box plots, and density plots, outliers were identified for removal. A p-value of less than 0.05 was considered as statistically significant.

## RESULTS

LVEF at baseline was 50.4±8.7% (range 34 – 65%). Follow-up at 6 months (FU 1) was available in 38/41 patients, with 21/38 subjects achieving LVEF improvement (P=0.0004 vs baseline; **Supplementary Table 3**). On univariate analysis, baseline LVEF (Odds ratio (OR) 0.80, 95% CI 0.68 – 0.90, P=0.002) and LN uptake (OR 0.21, 95% CI 0.10 – 0.79, P=0.03) reached significance **(Figure 1)**, indicating that lower LVEF and LN uptake were associated with improved outcome. Other PET parameters reflecting lymphatic organ uptake (spleen, OR 1.97, 95% CI 0.91 – 4.73, P=0.09; bone, OR 2.08, 95% CI 0.82 – 6.04, P=0.14) and infarct uptake (OR 0.39, 95% CI 0.10 – 1.38, P=0.15) failed in univariate analysis. We then created a multivariate Cox proportional hazards model including significant parameters from univariate analysis, along with established clinical parameters of systemic inflammation (WBC, CRP) and infarct size (LGE/mass) as morphological marker of myocardial damage. Baseline LVEF predicted functional recovery independently (OR 0.72, 95% CI 0.54 – 0.88, P=0.007), while LN uptake failed to reach significance (OR 0.28, 95% CI 0.02 – 2.07, P=0.24). Time-point of imaging serving as moderating variable also achieved no significance (OR 1.77, 95% CI 0.23 – 17.54, P=0.59), indicating that timing of PET (late vs. early imaging group) had no impact on functional recovery six months post-AMI. **Table 1** provides an overview of uni- and multivariate Cox regressions at FU 1.

**Table 1.**
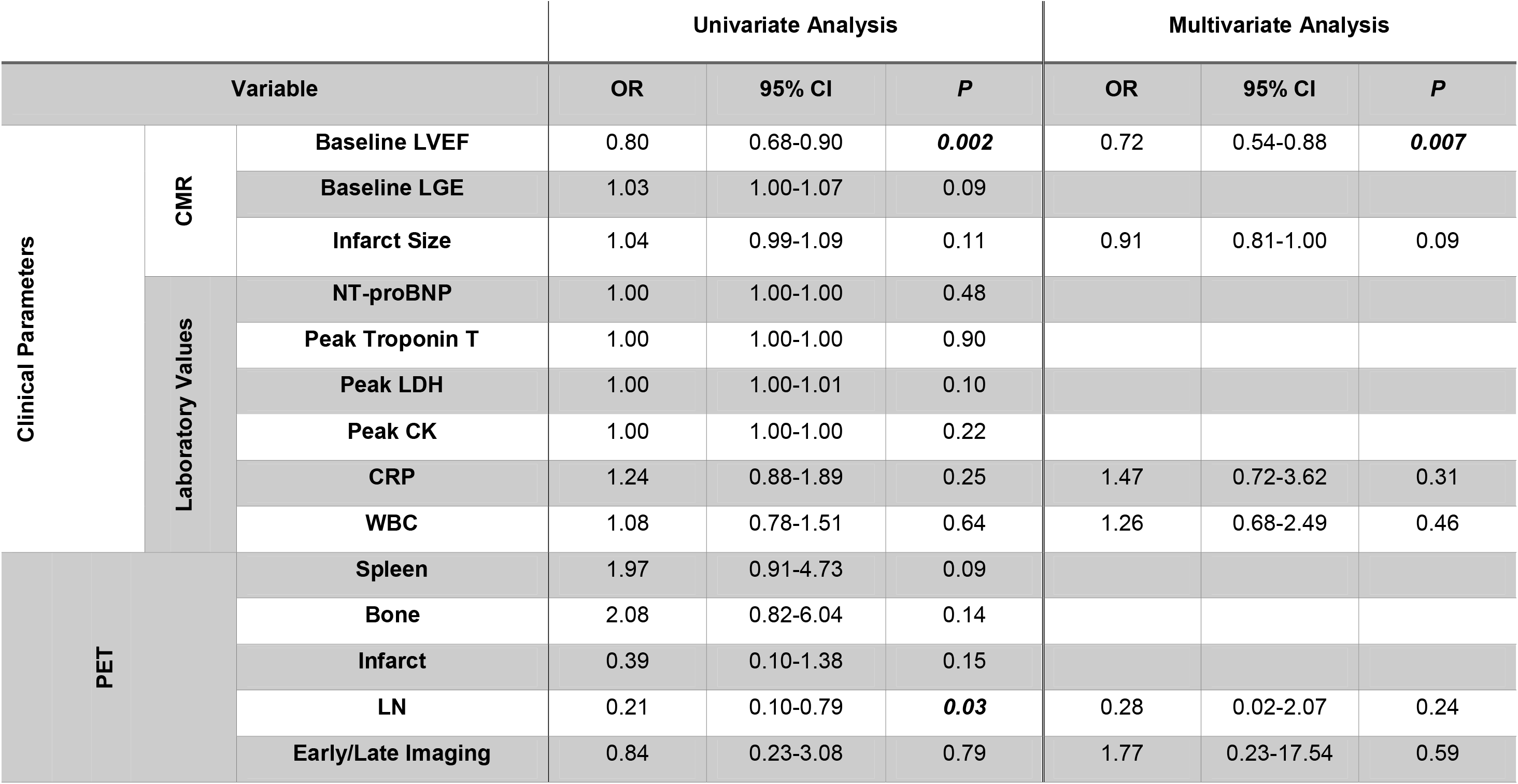
Uni-and multivariate Cox regression analysis six months after acute myocardial infarction (follow-up 1). OR=odds ratio, CI=confidence interval, LVEF=left ventricular ejection fraction, LGE=late gadolinium enhancement, NT-proBNP=N-terminal prohormone of brain natriuretic peptide, LDH=lactate dehydrogenase, CK=creatine kinase, CRP=C-reactive protein, WBC=white blood cell count, LN=lymph node. Infarct size reflected by LGE/mass (%). Early/late imaging refers to the time-point of ^68^Ga-PentixaFor PET/CT, including an early (2-4 days) or late imaging group (5-8 d) based on tracer availability. Significant values are marked in italic and bold.

**Figure 1.**
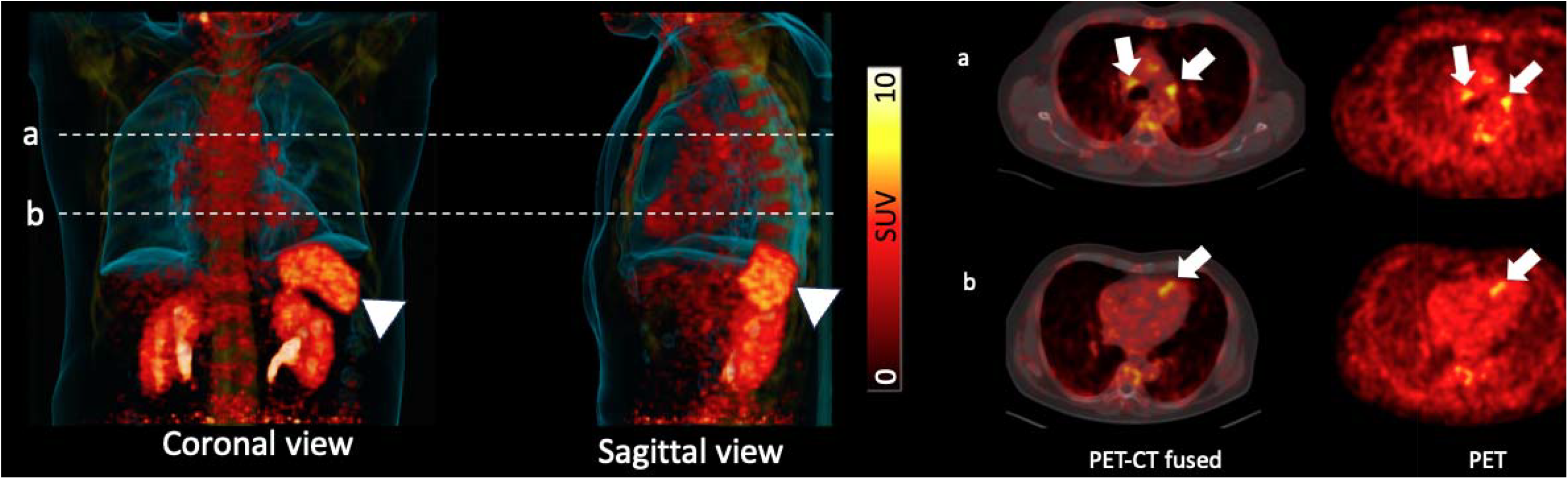
CXCR4-targeted ^68^Ga-PentixaFor PET/CT in a patient after acute myocardial infarction. Dotted lines on the PET/CT maximum intensity projections (MIP, left) in a coronal and sagittal view indicate trans-axial slides on the right. On trans-axial PET/CT and PET in (a), an intense PET signal can be observed in mediastinal lymph nodes (arrows), while in (b), the infarct area in the myocardium also exhibits intense in-vivo CXCR4 expression (arrows). On the MIPs (left), radiotracer signal can also be observed in the bone marrow and in the spleen (arrowheads). SUV=standardized uptake value.

At FU 2, follow-up was available in 36/41 patients, with 21/36 subjects achieving LVEF improvement (P=0.006 vs baseline; **Supplementary Table 3**). On univariate analysis, again baseline LVEF was significant for outcome 12 months post-AMI (OR 0.84, 95% CI 0.73 – 0.93, P=0.005), along with uptake in the spleen (OR 2.46, 95% CI 1.09 – 6.36, P=0.04, indicating that lower LVEF and increased splenic PET signal are associated with improved outcome. Other lymphatic organ uptake (bone, OR 2.29, 95% CI 0.86 – 7.31, P=0.12; LN, OR 0.39, 95% CI 0.09 – 1.42, P=0.17) and infarct uptake (OR 0.50, 95% CI, 0.13 – 1.80, P=0.29) failed to reach significance. We then applied the afore-mentioned multivariate Cox proportional hazards model with significant parameters from univariate analysis and established clinical parameters of inflammation and cardiac damage. Both baseline LVEF (OR 0.73, 95% CI 0.52 – 0.90, P=0.01) and uptake in the spleen (OR 4.17, 95% CI 1.21 – 22.35, P=0.04) remained significant for outcome at FU 2, indicating that lower baseline LVEF and increased splenic uptake can identify patients predisposed to long-term functional recovery. Again, imaging time-point of PET (late vs. early group) failed to reach significance in this model (OR 4.2, 95% CI 0.52 – 65.23, P=0.22), supporting the notion that scheduling of molecular imaging has no impact on long-term cardiac recovery. Uni- and multivariate Cox regressions for FU 2 are presented in **Table 2. Figure 2** shows two patients with low **(A)** and high **(B)** splenic uptake at CXCR4-directed PET/CT at baseline, with respective recovery and no recovery of LVEF at FU2.

**Table 2.**
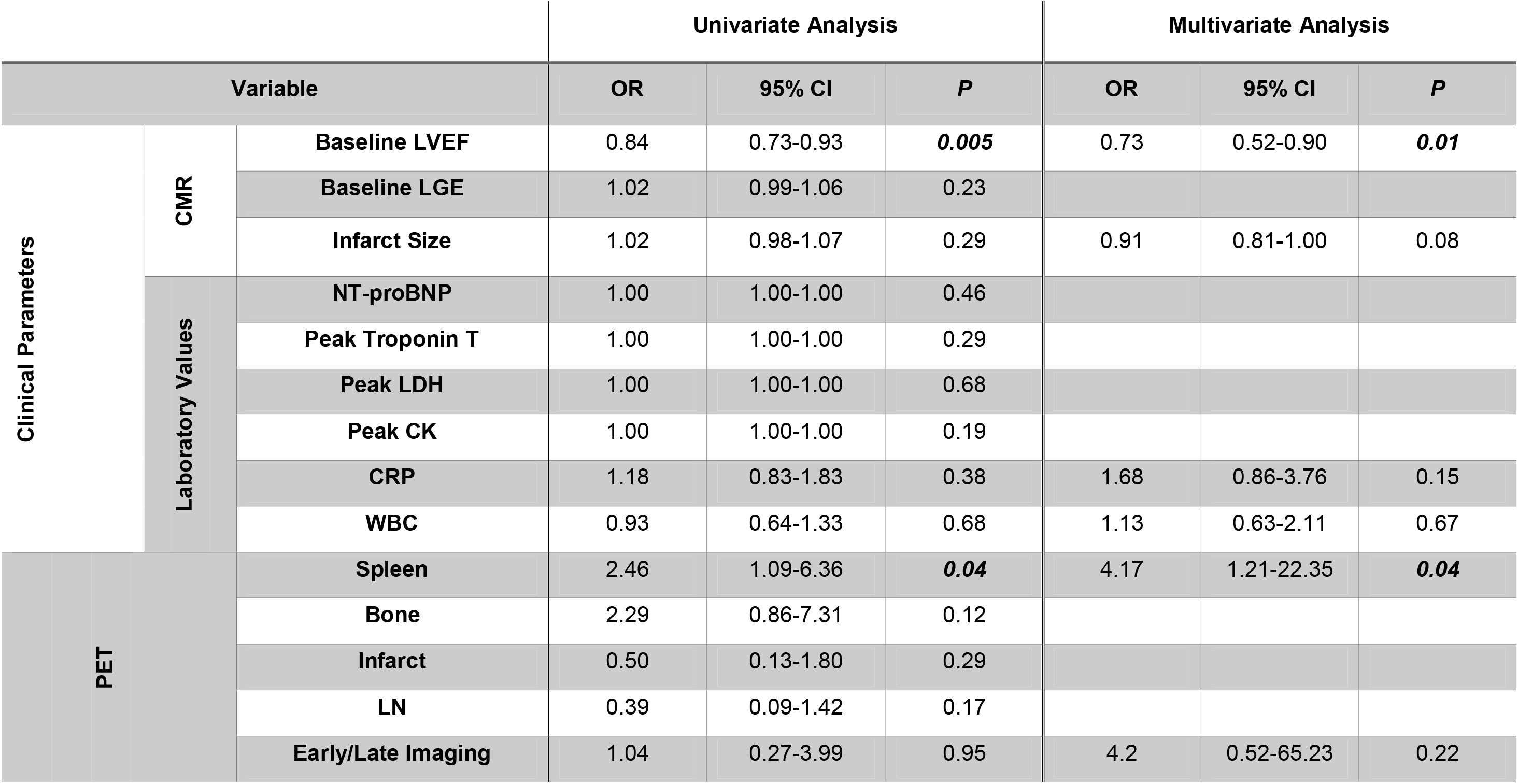
Uni- and multivariate Cox regression analysis twelve months after acute myocardial infarction (follow-up 2). OR=odds ratio, CI=confidence interval, LVEF=left ventricular ejection fraction, LGE=late gadolinium enhancement, NT-proBNP=N-terminal prohormone of brain natriuretic peptide, LDH=lactate dehydrogenase, CK=creatine kinase, CRP=C-reactive protein, WBC=white blood cell count, LN=lymph node. Infarct size reflected by LGE/mass (%). Early/late imaging refers to the time-point of ^68^Ga-PentixaFor PET/CT, including an early (2-4 days) or late imaging group (5-8 d) based on tracer availability. Significant values are marked in italic and bold.

**Figure 2.**
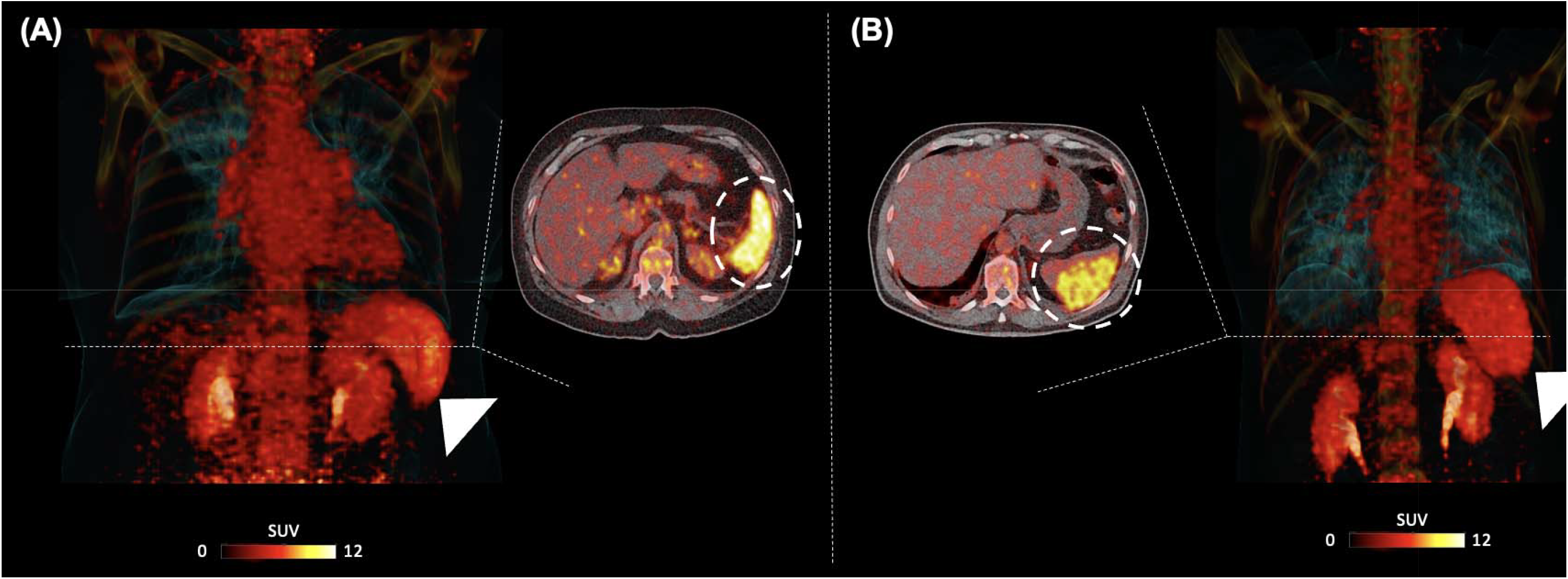
Maximum intensity projections and trans-axial PET/CT of the spleen (dotted circles, arrowheads) of patients with (A) and without (B) cardiac recovery twelve months after myocardial infarction. Patient on the left presented with high splenic tracer uptake and low baseline left ventricular ejection fraction and showed functional recovery. Patient on the right presented with low radiotracer uptake in the spleen and high baseline ejection fraction and presented with declining ejection fraction upon second follow-up. SUV=standardized uptake value.

## DISCUSSION

In timely re-perfused, ST-elevation AMI patients, the CXCR4-directed PET agent ^68^Ga-PentixaFor provided a non-invasive read-out of increased target expression in organs of systemic immune response. Established biomarkers of cardiac damage (infarct size) and systemic inflammation (CRP, WBC) failed to predict myocardial functional recovery. By contrast, cardiac baseline function and CXCR4 uptake in the spleen identified patients predisposed to functional recovery 12 months after AMI, indicating that our molecular imaging-derived biomarker provided complementary prognostic value for long-term cardiac outcome. These findings confirm the usefulness of a whole-body inflammatory-targeted molecular imaging approach focusing on organs of the systemic immune response.

In a recent study, CXCR4 PET signal in the infarct territory was linked to the occurrence of major cardiovascular events ^15^. Results of the present study go beyond the myocardium as the target region, as the CXCR4 PET signal in extra-cardiac lymphatic organs (LN, spleen) was associated with left ventricular recovery after AMI. Thus, we herein demonstrate an added clinical value of a whole-body PET approach also imaging distant organs involved in adaptive immunity post-AMI. In this regard, as shown in uni- and multivariate analyses, increased chemokine receptor PET signal in the spleen was tightly linked to long-term functional recovery 12 months after the acute event. Previous work has demonstrated that modulating the CXCR4 / C-X-C motif chemokine ligand 12 axis can deploy splenic regulatory T-cells from the spleen to the infarcted myocardium, which contribute to cardiac repair ^18,19^. With organs expressing CXCR4 in different cell types including lymphocytes, progenitor cells or natural killer cells^9-11^, our molecular imaging approach may therefore provide a non-invasive read-out of the inflammatory cell tracking between the primary site of damage (myocardium) and remote organs of hematopoietic activation (spleen).

There is increasing evidence of substantial benefits of immunomodulating drugs including colchicine or canakinumab for patients after MI ^1,3^, while treatment selection was based on systemic biomarkers of inflammation including CRP ^3^. While these studies aimed to reduce future vascular events, no specific biomarker is available for identifying patients that might benefit from immunological therapy to improve myocardial function. The observed value of the splenic PET signal for identifying patients predisposed to functional recovery and previous results on CXCR4 expression in the infarct territory for MACE prediction may open avenues for guiding cardiac repair by non-invasive molecular imaging ^15^. For instance, recent years have witnessed an increased use of highly specific, targeted drugs such as *in-vivo* re-programmed chimeric antigen receptor (CAR) T-cells to enhance cardiac repair, which also accumulate in the spleen to provide a target antigen reservoir ^20^. Those highly costly targeted interventions necessitate novel approaches for identifying subsets of patients that will most likely benefit from treatment, preferably by providing information on a (sub)cellular level about the absence or presence of the drug target in the myocardium or distant organs, e.g., target antigens in the spleen. Molecular imaging has multiple advantages relative to biopsies that are prone to sampling errors or blood-based systemic biomarkers which cannot provide segregated information on a local cardiac tissue level or on remote lymphatic organs ^5^. Relative to morphological imaging, PET can also determine the extent of the underlying pathophysiology, while CMR primarily focus on functional parameters ^7^. Nonetheless, in the present study, CMR-derived baseline LVEF and the splenic PET signal were relevant outcome predictors for functional recovery, thereby highlighting the importance of simultaneous assessments of morphological and molecular information for precisely identifying patients with best outcome. Thus, combined devices such as PET/MRI may therefore allow to determine those relevant parameters in a “one-stop-shop” setting, thereby facilitating a broader clinical use of the herein derived findings ^21^.

Our study has several limitations, including its small sample size in a single-center setting, with moderately impaired cardiac function at baseline. However, we still observed substantially decreased LVEF with a lower range of 34%. In this regard, imaging and therapeutic protocols including timely reperfusion therapies have been strictly followed, thereby presenting a cohort reflecting clinical reality. Second, due to logistic reasons including synthesis procedures and tracer availability, it was not feasible to conduct CXCR4 PET scans on the same day for every patient. Moreover, given the fact that chemokine receptors are overexpressed on a broad range of leukocytes, which are fluctuating over time early after AMI ^22^, it cannot be ruled out that such temporal dynamics may also interfere with severity of the PET signal conducted at different time-points. Nonetheless, with allocating our patients into an early (up to 4 days) and late imaging group (up to 8 days post-AMI), we observed no impact on outcome at FU 1 and FU 2 by using the time-point of PET imaging as a moderating variable.

In conclusion, in timely re-perfused, ST-elevation AMI patients, morphological and molecular imaging using chemokine receptor PET identified patients that are predisposed to functional recovery twelve months after the acute event. In this regard, increased splenic PET signal and low baseline cardiac function determined patients with LVEF improvement over 12 months, indicating that molecular imaging provides complementary prognostic value for long-term cardiac outcome. These findings confirm the usefulness of a whole-body inflammatory-targeted imaging approach focusing on the systemic immune response. In addition, our findings may open avenues for image-guided reparative interventions by identifying individuals that most likely benefit from targeted therapy based on PET signal strength in lymphatic organs. Our results also highlight the synergistic effects of both morphological and molecular imaging to identify AMI patients with later functional improvement, ideally by simultaneous assessments using PET/MRI.

## Data Availability

All data produced in the present study are available upon reasonable request to the authors

## Nonstandard Abbreviations and Acronyms

WBC: white blood cell count
CRP: c-reactive protein
CMR: cardiac magnetic resonance imaging
CXCR4: C-X-C motif chemokine receptor 4
AMI: acute myocardial infarction
LN: lymph nodes
PET/CT: positron emission tomography / computed tomography
FU: follow-up
CK: creatine kinase
LDH: lactate dehydrogenase
NT-proBNP: N-terminal prohormone of brain natriuretic peptide
SUV: standardized uptake values
MRI: magnetic resonance imaging
LGE: late gadolinium enhancement
LVEF: left ventricular ejection fraction

## Acknowledgements

We thank Dr. Dirk Mügge, Adelebsen, for statistical advice.

## SUPPLEMENTARY TABLES

**Supplementary Table 1.**
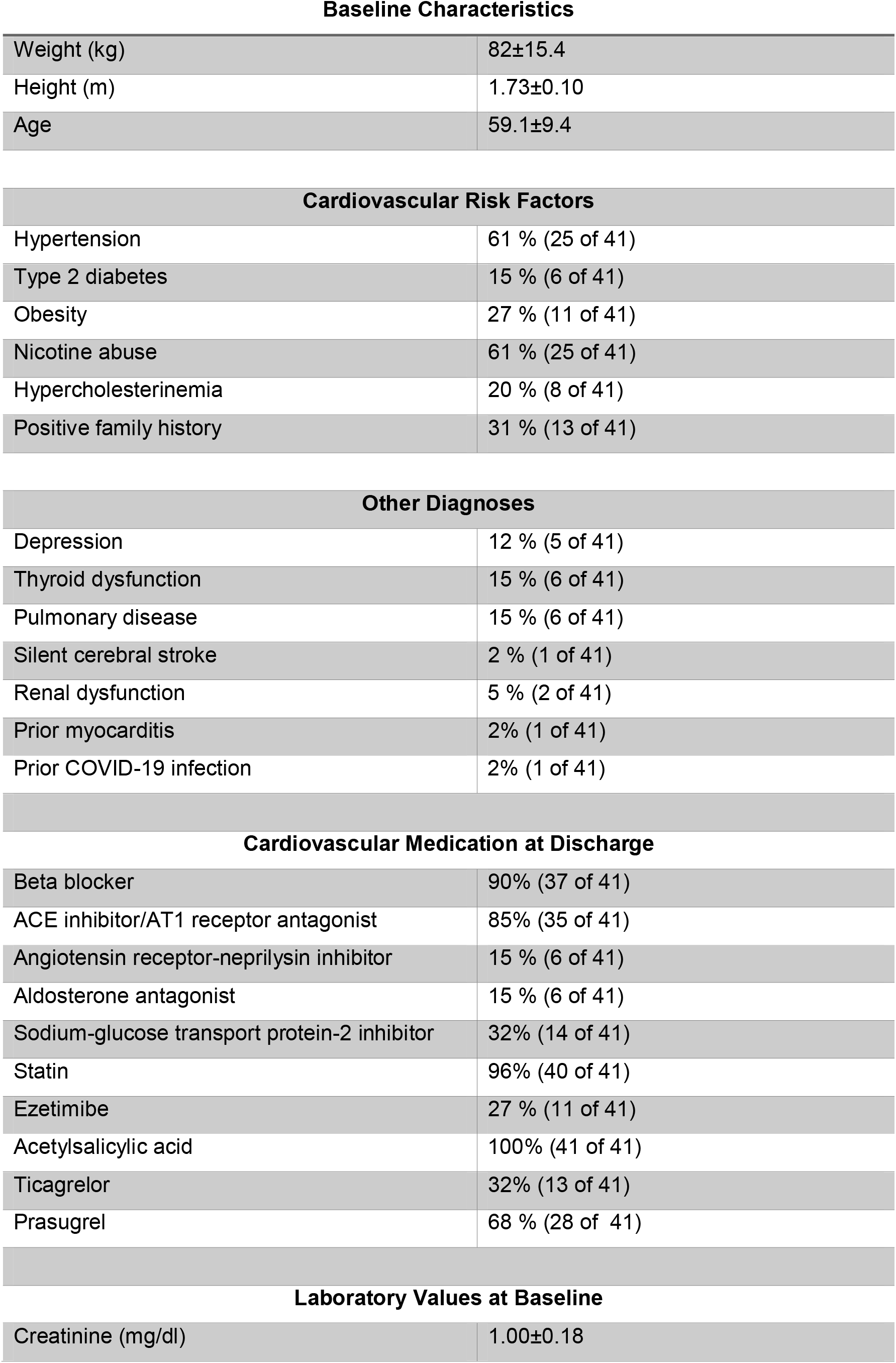
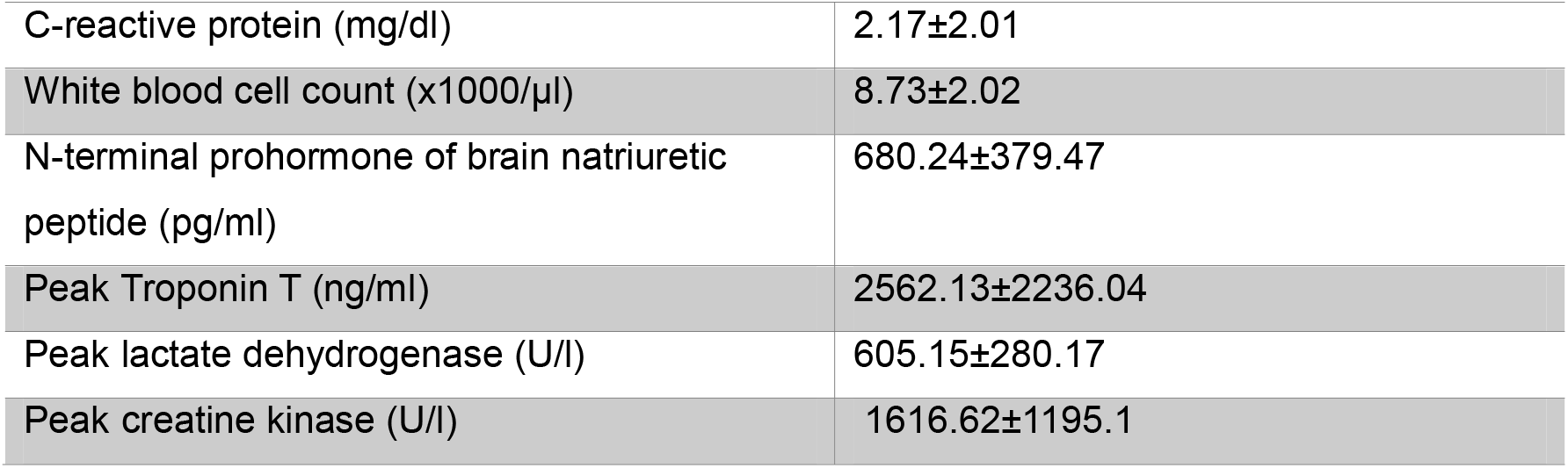
Patient’s characteristics at baseline.

Baseline Characteristics
|  |  |
| --- | --- |
| Weight (kg) | 82±15.4 |
| Height (m) | 1.73±0.10 |
| Age | 59.1±9.4 |
| Cardiovascular Risk Factors |  |
| Hypertension | 61 % (25 of 41) |
| Type 2 diabetes | 15 % (6 of 41) |
| Obesity | 27 % (11 of 41) |
| Nicotine abuse | 61 % (25 of 41) |
| Hypercholesterinemia | 20 % (8 of 41) |
| Positive family history | 31 % (13 of 41) |
| Other Diagnoses |  |
| Depression | 12 % (5 of 41) |
| Thyroid dysfunction | 15 % (6 of 41) |
| Pulmonary disease | 15 % (6 of 41) |
| Silent cerebral stroke | 2 % (1 of 41) |
| Renal dysfunction | 5 % (2 of 41) |
| Prior myocarditis | 2% (1 of 41) |
| Prior COVID-19 infection | 2% (1 of 41) |
| Cardiovascular Medication at Discharge |  |
| Beta blocker | 90% (37 of 41) |
| ACE inhibitor/AT1 receptor antagonist | 85% (35 of 41) |
| Angiotensin receptor-neprilysin inhibitor | 15 % (6 of 41) |
| Aldosterone antagonist | 15 % (6 of 41) |
| Sodium-glucose transport protein-2 inhibitor | 32% (14 of 41) |
| Statin | 96% (40 of 41) |
| Ezetimibe | 27 % (11 of 41) |
| Acetylsalicylic acid | 100% (41 of 41) |
| Ticagrelor | 32% (13 of 41) |
| Prasugrel | 68 % (28 of 41) |
| Creatinine (mg/dl) | 1.00±0.18 |
| C-reactive protein (mg/dl) | 2.17±2.01 |
| White blood cell count (x1000/μl) | 8.73±2.02 |
| N-terminal prohormone of brain natriuretic peptide (pg/ml) | 680.24±379.47 |
| Peak Troponin T (ng/ml) | 2562.13±2236.04 |
| Peak lactate dehydrogenase (U/l) | 605.15±280.17 |
| Peak creatine kinase (U/l) | 1616.62±1195.1 |

**Supplementary Table 2.**
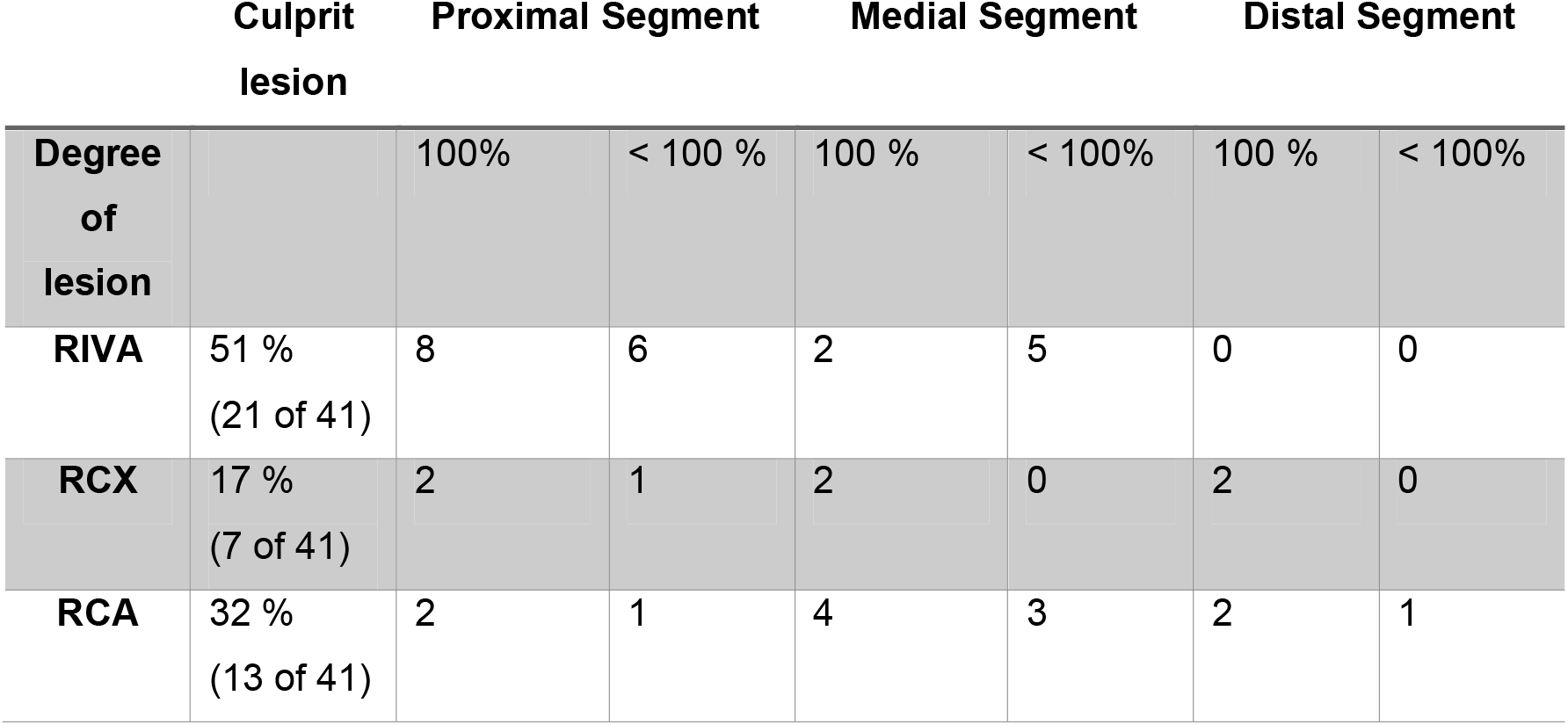
Culprit lesion and degree of stenosis. RIVA=Ramus interventricularis anterior. RCX=Ramus circumflexus. RCA=right coronary artery.

**Supplementary Table 3.**
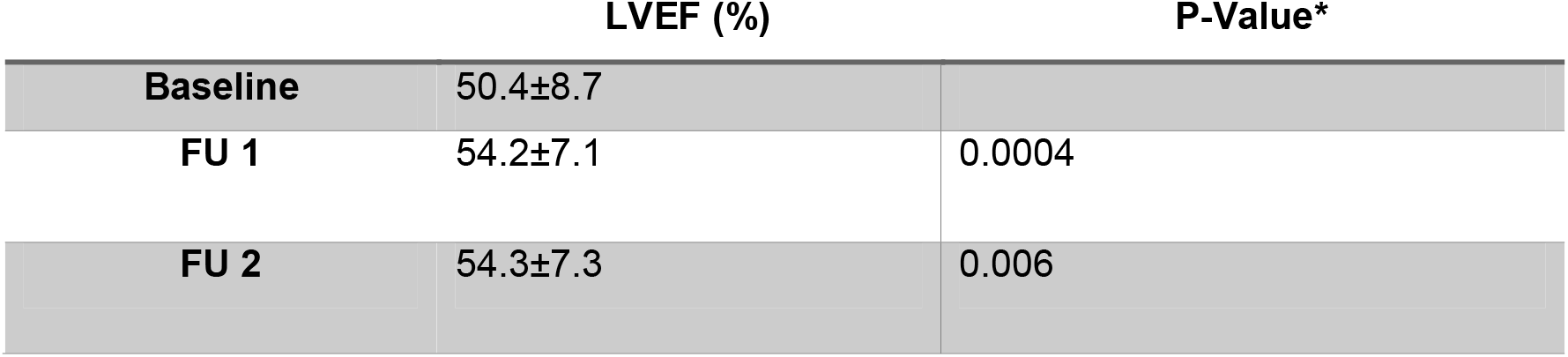
Left ventricular ejection fraction (LVEF) at baseline, follow-up (FU) 1 and FU 2. LVEF was assessed using cardiac magnetic resonance (CMR) at baseline after myocardial infarction, followed by repeated CMR at six (FU 1) and twelve months (FU 2). *refers to comparison with baseline.

|  | **LVEF (%)** | **P-Value\*** |
| --- | --- | --- |
| **Baseline** | 50.4±8.7 |  |
| **FU 1** | 54.2±7.1 | 0.0004 |
| **FU 2** | 54.3±7.3 | 0.006 |

